# Imprecision nutrition? Duplicate meals result in unreliable individual glycemic responses measured by continuous glucose monitors across four dietary patterns in adults without diabetes

**DOI:** 10.1101/2023.06.14.23291406

**Authors:** Aaron Hengist, Jude Anthony Ong, Katherine McNeel, Juen Guo, Kevin D Hall

## Abstract

**Background:** Continuous glucose monitors (CGMs) are being used to characterize postprandial glycemic responses and thereby provide personalized dietary advice to minimize glycemic excursions. However, the efficacy of such advice depends on reliable CGM responses.

**Objective:** To explore within-subject variability of CGM responses to duplicate meals in an inpatient setting.

**Methods:** CGM data were collected in two controlled feeding studies (NCT03407053 and NCT03878108) in 30 participants without diabetes capturing 1056 meal responses in duplicate ∼1 week apart from four dietary patterns. One study used two different CGMs (Abbott Freestyle Libre Pro and Dexcom G4 Platinum) whereas the other study used only Dexcom. We calculated the incremental area under the curve (iAUC) for each 2-h post-meal period and compared within-subject iAUCs using the same CGM for the duplicate meals using linear correlations, intra-class correlation coefficients (ICC), Bland-Altman analyses, and compared individual variability of glycemic responses to duplicate meals versus different meals using standard deviations (SDs).

**Results:** There were weak to moderate positive linear correlations between within-subject iAUCs for duplicate meals (Abbott r=0.47, p<0.0001, Dexcom r=0.43, p<0.0001), with low within-participant reliability indicated by ICC (Abbott 0.31, Dexcom 0.14). Bland-Altman analyses indicated wide limits of agreement (Abbott -31.3 to 31.5 mg/dL, Dexcom -30.8 to 30.4 mg/dL) but no significant bias of mean iAUCs for duplicate meals (Abbott 0.1 mg/dL, Dexcom -0.2 mg/dL). Individual variability of glycemic responses to duplicate meals was similar to that of different meals evaluated each diet week for both Abbott (SD_duplicate_ = 10.7 mg/dL, SD_week 1_ =12.4 mg/dL, SD_week 2_ =11.6 mg/dL, *p*=0.38) and Dexcom (SD_duplicate_ = 11.1 mg/dL, SD_week 1_ = 11.5 mg/dL, SD_week 2_ =11.9 mg/dL, *p*=0.60).

**Conclusions:** Individual postprandial CGM responses to duplicate meals were unreliable in adults without diabetes. Personalized diet advice based on CGM measurements in adults without diabetes requires more reliable methods involving aggregated repeated measurements.

This secondary analysis contains data from two trials registered at clinicaltrials.gov (NCT03407053 and NCT03878108).

## Introduction

Postprandial glycemic responses to different foods as measured by continuous glucose monitors (CGM) are highly variable between individuals, with some people exhibiting large glycemic excursions in response to one food compared to another, whereas a different person might experience the opposite results (1, 2). Such observations provide the rationale for personalizing diet advice to minimize glycemia excursions by attempting to identify the foods that result in reliably low postprandial glucose in each person (2, 3). The fundamental assumption of precision dietary advice is that repeated glycemic responses to the same meal within an individual are much less variable than their responses to different meals. However, this assumption has not been rigorously tested.

We investigated the reliability of within-subject postprandial CGM responses to duplicate ad libitum meals consumed ∼1 week apart by 15 participants residing at the NIH Clinical Center Metabolic Clinical Research Unit during two inpatient controlled feeding studies whose primary results have been reported elsewhere (4, 5). Study participants were presented with three daily meals from 7-day rotating menus for two weeks each such that each meal was provided in duplicate. Duplicate meals were included from four distinct dietary patterns. One was a minimally processed plant-based, low-fat diet, another was a minimally processed animal-based very-low carbohydrate ketogenic diet. The remaining two patterns had moderate macronutrient compositions, but one was high in ultra-processed foods, while the other was rich in unprocessed foods.

## Methods

We performed an exploratory analysis of data from two clinical research protocols approved by the institutional review board of the National Institute of Diabetes and Digestive and Kidney Diseases and are registered at clinicaltrials.gov (NCT03407053 and NCT03878108). Participants provided written informed consent and eligibility criteria for both studies were (1) ages 18–50 years; (2) body mass index >18.5 kg/m^2^; and (3) weight stable (<5% change in past 6 months). Both studies were within-subject, random-order crossover designs where participants were exposed to two diets for 14 days each on 7- day rotating menus, consuming each meal twice (once during week 1 and once during week 2). This enabled comparison of up to 21 repeated meals within each of the 4 dietary patterns. The daily menus had three meals (breakfast, lunch, and dinner), where the order within day was always fixed and the time of day when each meal was consumed was similar between the first and second week. Furthermore, the sequence of the daily menu was the same on the first and second week except in cases when the respiratory chamber day (whose menu was fixed within each diet pattern) had to be scheduled on a different day due to availability.

Meals were provided to participants alone in their inpatient rooms and photographs of the meals have been published previously alongside the primary outcomes (4, 5). Participants were instructed eat as much or as little food as they wanted and asked to not intentionally change their weight throughout the study. All foods were weighed to the nearest 0.1 g before and after consumption, and energy intake was calculated using ProNutra software (v.3.4, Viocare). A limitation of these studies with respect to our analyses of post-meal glycemic responses is that they included bottled water and snacks available throughout the day, but the timing of their consumption was not recorded.

Interstitial glucose concentrations were obtained from two brands of monitor: Abbott Freestyle Libre Pro (Abbott) and Dexcom G4 Platinum (Dexcom). In NCT03407053, some participants wore both Abbott and Dexcom, and in NCT03878108 study participants wore Dexcom. The Abbott device records glucose every 15 minutes and the Dexcom every 5 minutes. For accurate postprandial analysis, only duplicate meals with measured start time and sufficient data availability were included. For Abbott the mean (range) of duplicate meals within-participant was 28 (11 to 34) from 14 participants providing 392 total comparisons and for Dexcom the mean (range) of duplicate meals within-participant was 22 (2 to 49) from 30 participants providing 664 total comparisons. Data were aligned to the nearest 15-minute (Abbott) or 5-minute (Dexcom) CGM reading for calculation of post-meal responses. Baseline was assigned as the first time-point after the meal was provided. The 2-hour postprandial glucose incremental area under the curve (iAUC) was calculated for each meal using the trapezoid method, with dips below baseline assigned a negative value for iAUC (i.e., netAUC from (6)). Values of iAUC were reported as time-averaged glucose concentrations across the 2-h postprandial period.

Statistical analyses and data visualization were performed in R (v4.2.3) and GraphPad Prism (v9.5.0). Standard major axis regression was used to plot trends between meal 1 and meal 2 using lmodel2 in R. Simple linear correlation was calculated using Pearson’s correlation coefficient (r), with <0.4 interpreted as weak, 0.4 to 0.8 interpreted as moderate, and >0.8 interpreted as strong correlation. Repeatability was estimated by calculating the intra-class correlation coefficient (ICC) for glucose iAUCs. ICC was calculated using the following formula: ICC = participant variance / (participant variance + residual variance), which was generated from a linear mixed effects model with participant and residual error as random effects and meal and eating occasion as fixed effects (7). ICC values below 0.5 are considered as indicating poor reliability between measures (8). Bland-Altman analyses were conducted accounting for multiple observations per individual (9).

To examine individual glycemic variability in response to duplicate meals as compared with different meals, we partitioned the total iAUC variance between diet pattern, meal type (breakfast, lunch, dinner), menu day, and duplicate meals between successive weeks to compute the standard deviation (SD) of duplicate iAUC responses as follows: 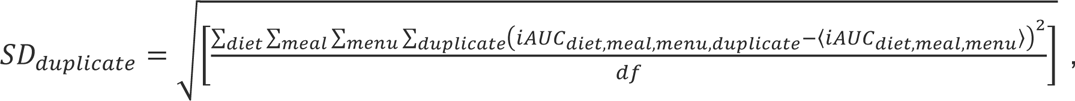 where df is the degrees of freedom and <iAUC_diet,meal,menu_> is the average iAUC over duplicate meals. Similarly, we computed individual SDs of the iAUC responses to different meals on each week of 7-day rotating menus as follows:

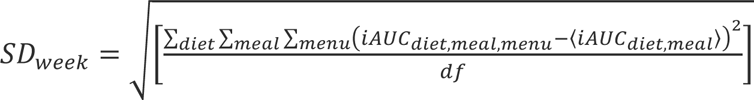

where df is the degrees of freedom and <iAUC_diet,meal_> is the average iAUC over the 7-day week of menus. One-way ANOVA with Bonferroni correction was used to compare SDs of week 1, week 2, and duplicate meal responses. Energy intake of meals in week 1 and week 2 were compared using a paired t-test. Significance was accepted as *p*≤0.05.

During the inpatient stay of each study, venous glucose measurements were obtained during oral glucose tolerance tests (OGTTs) in both studies and mixed meal tolerance tests (MMTTs) in NCT03878108 (Dexcom only) whilst wearing CGMs. This allowed for an assessment of individual variability between simultaneously measured venous and CGM determined iAUCs in response to OGTTs and MMTTs to provide an index of how much of the CGM iAUC variability in response to duplicate meals might be due to variability in the iAUC determined by CGM as compared to simultaneous venous measurements. So, we evaluated the SD of the difference between CGM and venous iAUCs in response to the same meal test and compared this to the SD of the duplicate CGM iAUC responses.

To potentially identify predictors of individual variability in postprandial glucose response to duplicate meals, we used forward stepwise linear regression to estimate the contribution of measured behavioral variables using the stepAIC function of the R package ‘MASS’. The response variable was difference between duplicate meal postprandial glucose responses assessed as either iAUC or total area under the curve (tAUC). Predictor variables included differences in baseline blood glucose (for iAUC only), difference in time taken to consume the meals, number of days between meals, difference in energy intake from snack consumption, type of meal (breakfast, lunch, or dinner), difference in consumed meal-specific macronutrients energy (protein, fat, and carbohydrate), presence of exercise ≤ 30 minutes before the start of a meal, and presence of exercise during the postprandial period. During the study, participants were instructed to complete three 20-minute light-to-moderate intensity exercise sessions on a bicycle ergometer with standardized wattage and speed (between 30-40% heart rate reserve). Meal duplicates with inaccurate meal or exercise timing were omitted from the regression analyses.

## Results

### Glycemic responses to the same meals eaten on separate occasions are unreliable

We investigated 30 participants whose characteristics are shown in **Table 1**, who were presented with duplicate meals on two consecutive weeks exactly 7 days apart for 85% of Abbott measurements and 63% of Dexcom measurements. **Figure 1A** plots the iAUC responses to meals consumed on week 2 versus the duplicate meals on week 1 measured in the same participants using the Abbott device. **Figure 1B** plots analogous data obtained using the Dexcom device. Regardless of CGM, there were weak to moderate positive linear correlations between the within-subject iAUC responses to duplicate meals across all dietary patterns (Abbott r=0.47, *p*<0.0001, Dexcom r=0.43, *p*<0.0001). Linear correlations were similar when meals were split into breakfast, lunch, and dinner (Abbott breakfast r=0.41, lunch r=0.55, dinner r=0.44; Dexcom breakfast r=0.41, lunch r=0.43, dinner r=0.40, all *p*<0.0001).

**Figure 1.**
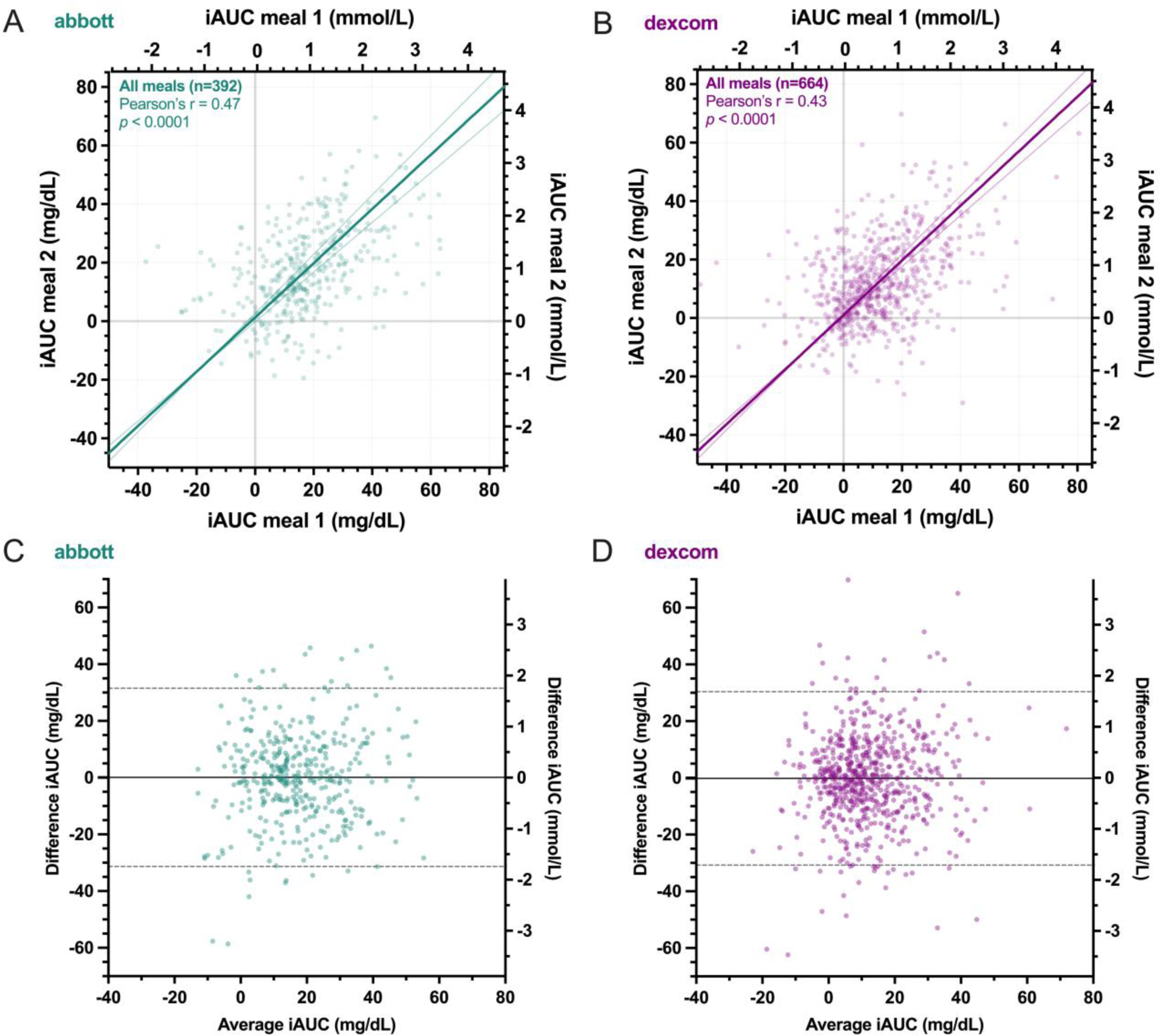
Comparison of incremental area under the curve (iAUC) of postprandial glucose responses to duplicate meals using A) Abbott and B) Dexcom continuous glucose monitors. Trendline is major axis regression with 97.5% CIs. Bland-Altman plots of the iAUC differences between duplicate meals versus the average of both measurements using C) Abbott and D) Dexcom devices. Solid line indicates mean bias and dashed lines indicate 95% limits of agreement.

**Table 1.** Baseline characteristics of participants included in analyses.

|  | Abbott<br>(n=14) | Dexcom<br>(n=30) |
| --- | --- | --- |
| Sex (F,M) | F=8, M=6 | F=15, M=15 |
| Age (y) | 31 ±8 | 30 ±7 |
| Body mass (kg) | 73.0 ±13.3 | 80.6 ±19.4 |
| Body mass index (kg/m<sup>2</sup>) | 25.5 ±5.2 | 27.6 ±6.3 |
| Body fat (%) | 28.9 ±11.1 | 31.9 ±9.9 |

Data mean ± SD.

Intra-class correlation coefficients (ICCs) were 0.31 for Abbott and 0.14 for Dexcom, indicating that there was a low tendency for glucose responses to be similar in duplicate meals in the same participant. Across all duplicate meals and subjects, Bland-Altman plots are shown in **Figures 1C** and **1D** indicating a low mean bias between iAUC responses to duplicate meals (Abbott 0.1 mg/dL, Dexcom -0.2 mg/dL), but there was a large variability indicated by the wide 95% limits of agreement (LoA) for both CGMs (Abbott -31.3 to 31.5 mg/dL, Dexcom -30.8 to 30.4 mg/dL).

**Supplemental Figure 1A** and **1B** plot the differences in glycemic responses to duplicate meals for each individual participant using the Abbott and Dexcom devices, respectively, and show highly variable glycemic responses when the same participant consumed duplicate meals on separate weeks, regardless of the CGM device. However, the iAUC bias was relatively low for most participants when averaged across different duplicate meals. **Supplemental Figure 1C** and **1D** plot the same data separated by individual duplicate meals as measured using the Abbott and Dexcom devices, respectively, and indicate highly variable individual glycemic responses to duplicate meals, regardless of the CGM device. Nevertheless, the iAUC bias was relatively low for most meals when averaged across participants.

### Similar individual glycemic response variability to duplicate versus different meals

Surprisingly, we found that everyone’s glycemic response variability to duplicate meals was similar to the variability in their glycemic responses to different meals. **Figure 2A** plots the SD of the glycemic responses in each individual participant to different meals eaten in either week 1 or week 2 along with the SD of their glycemic responses to duplicate meals for the Abbott device. **Figure 2B** plots analogous data from the Dexcom device. Regardless of device, the variability in the glycemic response to duplicate meals was similar to each participant’s glycemic response variability to different meals (Abbott: SD_week 1_ = 12.4 mg/dL, SD_week 2_ = 11.6 mg/dL, SD_duplicate_ = 10.7 mg/dL, *p*=0.38; Dexcom: SD_week 1_ = 11.5 mg/dL, SD_week 2_ = 11.9 mg/dL, SD_duplicate_ = 11.1 mg/dL, *p*=0.60).

**Figure 2.**
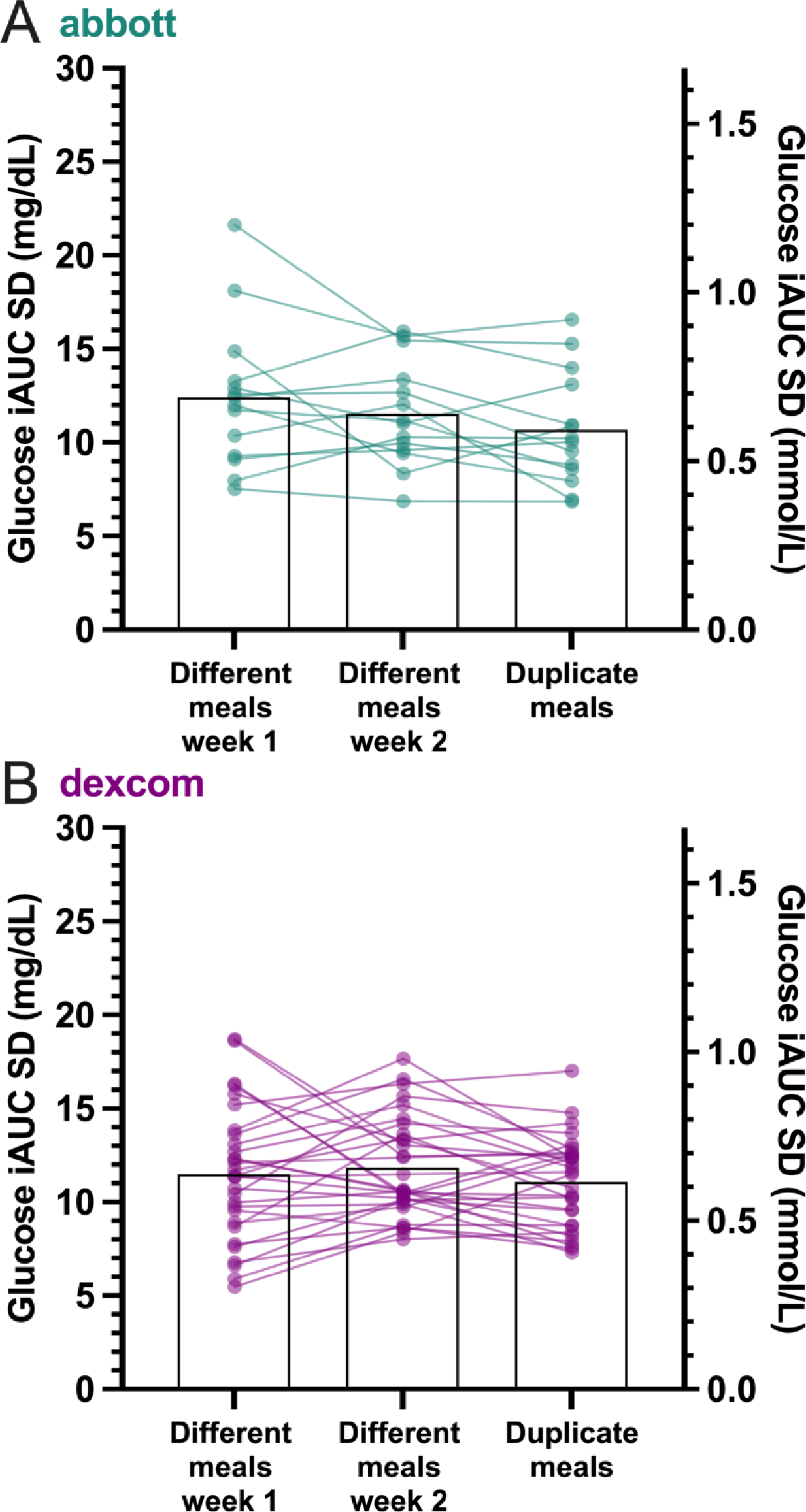
Mean and individual participant standard deviations (SD) of postprandial glucose responses between different meals across week 1, different meals across week 2, and duplicate meals between weeks using A) Abbott and B) Dexcom devices.

### Comparing venous and CGM-derived interstitial glucose responses to oral glucose and mixed meal tolerance tests

Mean ±SEM iAUC responses to OGTTs were similar between venous (39.5 ±3.8 mg/dL) and Abbott CGM (39.5 ±3.9 mg/dL, *p*=0.99; **Figure 3A**) and were moderately correlated (r=0.73, *p*<0.0001; **Figure 3B**). Mean ±SEM iAUC responses to OGTTs and diet-specific MMTTs were similar between venous (38.1 ±2.8 mg/dL) and Dexcom CGM (36.8 ±2.9 mg/dL, *p*=0.53; **Figure 3D**) and were also moderately correlated (r=0.72, *p*<0.0001; **Figure 3E**). Note that glycemic responses to very low carbohydrate meals were low for all participants. The SD of venous to CGM responses were not significantly different to duplicate meal SDs for Abbott (*p*=0.28; **Figure 3C**) and even higher for Dexcom (*p*=0.01; **Figure 3F**) suggesting that CGM imprecision of individual iAUC measurements contributes to the observed unreliable quantification of glycemic responses to meals.

**Figure 3.**
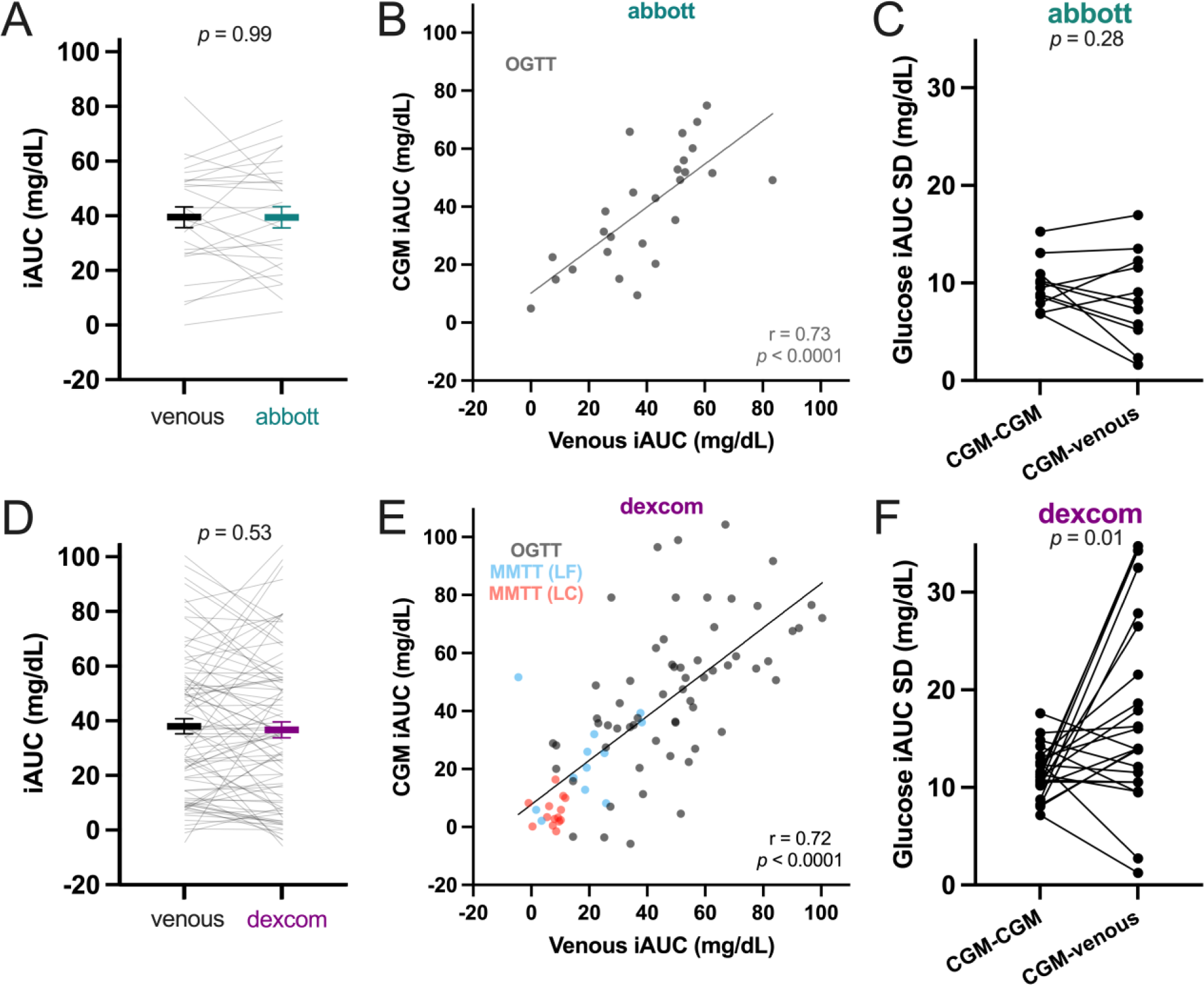
Mean ±SEM and individual participant comparisons of venous incremental area under the curve (iAUC) responses to A) oral glucose tolerance tests (OGTTs) with Abbott (n=26), and D) OGTTs and mixed meal tolerance tests (MMTT) with Dexcom (n=87). Linear regression of B) Abbott and E) Dexcom iAUC (Pearson’s r). Individual participant standard deviations (SD) of postprandial glucose responses to duplicate meals using CGM and venous to CGM comparisons for C) Abbott and F) Dexcom. Paired t-tests used for A,C,D,F.

### Potential factors affecting variability in glycemic response to duplicate meals

We explored numerous factors that may have contributed to explaining the glycemic response variability to duplicate meals. Firstly, there was a moderate negative linear correlation between differences in baseline glucose and differences in iAUC (Abbott r=- 0.42 *p*<0.0001, Dexcom r=-0.48 *p*<0.0001), suggesting baseline glucose concentrations may have contributed to the low repeatability of glucose iAUC. Repeating analyses with tAUC rather than incremental moderately increased ICC for both Abbott (0.57) and Dexcom (0.23).

Because food intake was ad libitum in our studies, we investigated whether differences in meal energy intake between duplicate meal weeks affected our findings. For Abbott, mean (95% CI) meal energy intake was 777 (746 to 808) kcal in week 1 and 744 (712 to 777) kcal in week 2 (*p*=0.0007). For Dexcom, mean (95% CI) meal energy intake was 790 (761 to 819) kcal in week 1 and 770 (742 to 798) kcal in week 2 (*p*=0.02). Energy intake between duplicate meals was strongly positively correlated (Abbott r=0.83 *p*<0.0001, Dexcom r=0.83 *p*<0.0001) and there was a weak positive correlation between differences in energy intake and differences in glucose iAUC (Abbott r=0.24 *p*<0.0001, Dexcom r=0.13 *p*=0.008). However, repeating our analyses using only duplicate meals where energy intake was within 100 kcal between meals did not materially affect our results regarding the iAUC correlations (Abbott n=201, r=0.43 *p*<0.0001, Dexcom n=314, r=0.45 *p*<0.0001), or ICC (Abbott 0.29, Dexcom 0.16), or Bland-Altman analyses (Abbott bias -0.5 mg/dL, LoA -31.9 to 30.9 mg/dL Dexcom bias -0.4 mg/dL, LoA -31.2 to 30.3 mg/dL). Using only meals where energy intake was within 100 kcal, the variability in the glycemic response to duplicate meals remained similar to each participant’s glycemic response variability to different meals (Abbott: SD_week 1_ = 12.4 mg/dL, SD_week 2_ = 12.0 mg/dL, SD_duplicate_ = 10.7 mg/dL, *p*=0.43; Dexcom: SD_week 1_ = 12.9 mg/dL, SD_week 2_ = 11.8 mg/dL, SD_duplicate_ = 11.4 mg/dL, *p*=0.45).

In addition to the three daily meals provided, participants were also given snacks that could be consumed at any time of day. To examine whether our results may have been affected by differences in snack intake between days with duplicate meals, we filtered the data such that snack intake was <200 kcal on both duplicate meal days resulting in 136 duplicates meals available for Abbott and 245 for Dexcom. For Abbott, mean (95% CI) meal energy intake was 791 (738 to 845) kcal in week 1 and 748 (694 to 802) kcal in week 2 (*p*=0.008) and mean (95%) snack intake was 39 (28 to 50) kcal/d in week 1 and 32 (23 to 40) kcal/d in week 2 (*p*=0.31). For Dexcom, mean (95% CI) meal energy intake was 720 (675 to 765) kcal in week 1 and 709 (667 to 752) kcal in week 2 (*p*=0.34) and mean (95%) snack intake was 29 (22 to 37) kcal/d in week 1 and 20 (14 to 25) kcal/d in week 2 (*p*=0.02). Repeating our analyses using only meals where snack intake was less than 200 kcal did not materially affect our results regarding the iAUC correlations (Abbott r=0.59 *p*<0.0001, Dexcom r=0.48 *p*<0.0001), or ICC (Abbott 0.34, Dexcom 0.19), or Bland-Altman analyses (Abbott bias 0.4 mg/dL, LoA -27.3 to 28.2 mg/dL; Dexcom bias 1.2 mg/dL, LoA -29.5 to 31.8 mg/dL). Using only meals where daily snack intake was less than 200 kcal, the variability in the glycemic response to duplicate meals remained similar to each participant’s glycemic response variability to different meals (Abbott: SD_week 1_ = 11.3 mg/dL, SD_week 2_ = 10.2 mg/dL, SD_duplicate_ = 8.8 mg/dL, *p*=0.30; Dexcom: SD_week 1_ = 10.9 mg/dL, SD_week 2_ = 9.5 mg/dL, SD_duplicate_ = 10.4 mg/dL, *p*=0.43).

### Explaining variance in response to duplicate meals using known behavioral variables

For Abbott, the difference in baseline glucose, carbohydrate content of meals, energy intake from consumption of snacks, and the time taken to consume meals were identified as predictor variables for the difference in iAUC between duplicate meals (**Table 2**). For tAUC, predictor variables were the difference in carbohydrate content and energy intake from snacks (**Table 2**). For Dexcom, the difference in baseline glucose, difference in carbohydrate content of meals, postprandial exercise occurring only in meal 1, postprandial exercise occurring only in meal 2, and difference in time to consume meals were identified as predictor variables for the difference in iAUC between duplicate meals (**Table 3**. For tAUC, the difference in carbohydrate content, presence of postprandial exercise only in meal 1, presence of postprandial exercise only in meal 2, and difference in energy intake from snacks were identified as predictor variables (**Table 3**). While these identified predictor variables significantly contributed to individual variability in postprandial glucose responses to duplicate meals, they explained only a small amount (≤ 25%) of the variability as determined by the coefficients of determination (R^2^) values in the resulting linear regression models (Tables 2 and 3).

**Table 2.** Forward stepwise linear regression of incremental (iAUC) and total area under the curve (tAUC) interstitial glucose responses to duplicate meals eaten on separate days using Abbott continuous glucose monitors (CGMs).

| *Variable* | **Difference in iAUC**<br>**(mg/dL)** |  |  | **Difference in tAUC**<br>**(mg/dL)** |  |  |
| --- | --- | --- | --- | --- | --- | --- |
|  | *Coefficient* | *Std.*<br>*Error* | *P-Value* | *Coefficient* | *Std.*<br>*Error* | *P-Value* |
| (Intercept) | -0.46 | 0.80 | <u>0.568</u> | -0.80 | 0.94 | 0.394 |
| Difference in Baseline Glucose (mg/dL) | -0.42 | 0.05 | **<0.001** |  |  |  |
| Difference in Carbohydrate Consumed (kcal) | 0.04 | 0.01 | **<0.001** | 0.02 | 0.01 | **0.013** |
| Difference in Snack Energy Intake (kcal) | -0.01 | 0.00 | **0.002** | -0.01 | 0.00 | **0.006** |
| Difference in Time to Consume Meal (min) | -0.13 | 0.06 | **0.030** |  |  |  |
| Observations | 333 |  |  | 333 |  |  |
| R<sup>2</sup> / R<sup>2</sup> adjusted | 0.247 / 0.238 |  |  | 0.040 / 0.034 |  |  |

**Table 3.** Forward stepwise linear regression of incremental (iAUC) and total area under the curve (tAUC) interstitial glucose responses to duplicate meals eaten on separate days using Dexcom continuous glucose monitors (CGMs).

| *Variable* | **Difference in iAUC**<br>**(mg/dL)** |  |  | **Difference in tAUC**<br>**(mg/dL)** |  |  |
| --- | --- | --- | --- | --- | --- | --- |
|  | *Coefficient* | *Std.*<br>*Error* | *P-Value* | *Coefficient* | *Std.*<br>*Error* | *P-Value* |
| (Intercept) | 1.16 | 0.69 | 0.094 | 2.15 | 0.88 | **0.015** |
| Difference in Baseline Glucose (mg/dL) | -0.40 | 0.03 | **<0.001** |  |  |  |
| Difference in Carbohydrate Consumed (kcal) | 0.02 | 0.01 | **<0.001** | 0.02 | 0.01 | **0.010** |
| Meal 1 Postprandial Exercise (Y/N) | -4.05 | 1.59 | **0.011** | -3.73 | 2.02 | 0.065 |
| Meal 2 Postprandial Exercise (Y/N) | 0.26 | 1.50 | 0.864 | 2.03 | 1.89 | 0.285 |
| Difference in Time to Consume Meal (min) | -0.08 | 0.05 | 0.117 |  |  |  |
| Difference in Snack Energy Intake (kcal) |  |  |  | 0.01 | 0.00 | **0.010** |
| Observations | 580 |  |  | 580 |  |  |
| R<sup>2</sup> / R<sup>2</sup> adjusted | 0.250 / 0.243 |  |  | 0.029 / 0.023 |  |  |

### Implications for Meal Ranking

Given the low reliability of iAUC responses, advice to eat meals having low postprandial glucose responses (i.e., bottom tertile) based on a single meal test does not necessarily result in low glycemic excursions in response to the same meals in the future. **Supplemental Figure 2** shows that meals in the bottom iAUC tertile on week 1 were 93% and 123% lower than the mean across all meals for Abbott and Dexcom, respectively. However, the same meals on week 2 were only 48% and 49% lower than average. Conversely, **Supplemental Figure 2** also shows that advice to avoid meals with elevated iAUC, in week 1 shows the inverse. Meals in the upper iAUC tertile on week 1 were 99% and 132% higher than the mean across all meals for Abbott and Dexcom, respectively. However, the same meals on week 2 were only 59% and 60% higher than average, suggesting regression to the mean with repeated measures.

## Discussion

CGM devices are becoming widely used in people without diabetes as part of commercial precision nutrition programs that provide personalized diet advice (10), however, CGM responses need to be precise and accurate to be useful (11). The fundamental assumption of personalized or “precision” nutrition is that an individual’s responses to repeated meals are less variable than their responses to different meals. Otherwise, it would be impossible to provide reliable advice to avoid meals that result in poor responses. Previous work found relatively reliable postprandial CGM responses to a small number of duplicate simple meals like bread (2) or muffins (1), but such meals are not representative of multicomponent meals that are the focus of personalized dietary advice in the real-world. Surprisingly, our study found that the reliability of postprandial CGM responses to many duplicate multicomponent meals was poor and that the within-subject variability to duplicate meals was roughly as large as the variability across different meals. Perhaps this is why recent randomized trials comparing personalized nutrition interventions focused on glycemic responses observed small effects for mean glucose (within 7 mg/dL, 0.39 mmol/L) and HbA1c (within 0.14%) (12), or no differences in glycemic variability and HbA1c (13) as compared to general diet advice.

We recently demonstrated that postprandial glycemic responses using two different brands of CGMs simultaneously worn on different anatomical locations resulted in only moderate correlations of within-subject postprandial responses to simultaneously measured multicomponent meals (r=0.68) and modest concordance of the meal rankings by iAUC (Kendal rank correlation = 0.43) (14). A subsequent study using simple test meals (i.e., muffins, milkshakes, and energy bars) confirmed that simultaneous within-subject postprandial iAUCs measured using different CGM devices were only moderately correlated (r=0.61) but the rank order of these simple meals according to iAUC was more concordant (Kendall rank correlation = 0.68) than the rankings of multicomponent meals in our previous study, perhaps reflecting the formulation of simple test foods to have wide differences in glycemic load (15). Interestingly, using identical CGMs in the same anatomical location resulted in much better agreement (r=0.97; Kendall rank correlation = 0.87) suggesting that a given CGM device provides valid measures of postprandial glycemic responses to simple test meals on a single occasion (15). However, this does not address the reliability of within-subject responses to repeated meals.

The relatively low within-subject reliability of postprandial CGM responses to duplicate multicomponent meals in our study occurred under highly controlled metabolic ward conditions where meal order within each day was standardized and was typically preceded by a previous standardized day. Whilst less reflective of free-living conditions, such inpatient controlled feeding studies reduce the amount of variability explained by behavioral factors, enabling better understanding of the amount of glycemic variability that can be explained by ingestion of meals, providing a better indication of measurement error (16, 17). However, despite the strengths of our inpatient controlled feeding design, our study had several limitations. First, the primary aims of the original studies were to measure ad libitum energy intake differences between dietary patterns, therefore duplicate meals were not necessarily consumed in identical amounts, although energy intake of duplicate meals was highly correlated and repeating the analyses using only meals within 100 kcal of each other did not change interpretation. Furthermore, despite the regimented meal order and timing achieved with implementation of the 7-day rotating menus, snacks were available for consumption at any time of day which may have differentially affected meal responses. Re-analysis using only duplicate meals on days when snack intake was below 200 kcal or when the energy intake difference between duplicate meals was <100 kcal did not materially affect our results.

The two strongest predictor variables of postprandial glucose differences in response to duplicate meals were differences in baseline glucose and carbohydrates consumed. Interestingly, the impact of exercise on postprandial glucose responses was only identified using Dexcom and not Abbott. The presence of a 20-minute bout of light-to-moderate intensity exercise during the postprandial period decreased glucose responses in the Dexcom, similar to previous findings (18). The discrepancy between CGMs could be due to the anatomical location of the monitors in proximity to physically active tissue (19), and increased subcutaneous adipose tissue blood flow (20). In our previous analysis, the discrepancy between Abbott (arm) and Dexcom (abdomen) was larger in in individuals with higher body fat (14). Regardless of CGM, the variation in postprandial glucose excursions to duplicate meals was only weakly correlated with the measured predictor variables thought to influence glucose responses in our regression model. This suggests it would be very difficult to explain the variance in CGM response between duplicate meals by collecting data on behavior in a free-living setting.

Repeating analyses with tAUC rather than iAUC modestly improved reliability for both Abbott and Dexcom. The moderate reliability of Abbott was similar to reliability previously reported for 4-h glucose tAUCs using venous samples from duplicate mixed meals, performed under standardized conditions twice within 28 days in adults with normal glucose tolerance (21), but ICC for Dexcom was still much lower when using tAUC.

Due to the inpatient setting, our study has limited generalizability to free-living people. However, free-living behaviors will likely further increase the within-subject variability of CGM responses to similar meals. A plethora of modifiable behavioral factors can also influence postprandial glycemic responses to the same meal within an individual and the reasons for the variable responses to repeated meals in our study are presently unknown. In our study, meals were ad libitum, but participants tended to eat similar amounts of the repeated meals and differences in energy intake did not seem to account for the differences in glycemic response, as the variability was similar when only analyzing meals with similar energy intake within 100 kcal (data not shown). However, variations in the sequence of foods consumed within the repeated multicomponent ad libitum meals may have contributed to the variability because food sequence has been previously shown to result in varying glycemic responses in people with and without type 2 diabetes (22-25).

Physical activity differences may have also played a role, as previous studies have shown that breaking up prolonged sitting with small amounts of physical activity during the postprandial period reduces postprandial glycemia (26-28), and even leg fidgeting may have an effect (29). Sleep quality and bedtime has recently been associated with changes in CGM-derived measures of postprandial glucose (30), so variations in sleep quality may have contributed to differences in the studies presented. Importantly, if such behavioral factors are indeed important contributors to meal glycemic responses, then an enormous amount of data may be required to capture these behavioral determinants and reliably predict an individual’s glucose excursions and thereby provide personalized “precision” diet advice.

Participants wore CGMs during standardized meal tests (either OGTTs or diet-specific MMTTs) concurrent with venous blood measures to provide some indication of the contribution of technical error to the observed variable postprandial responses, acknowledging there is a 5-6 minute lag time for glucose moving from intravascular to interstitial compartments (31). The correlation between venous and CGM was responses were similar to values recently reported, but we did not observe a mean difference between CGM and venous iAUCs, compared to the ∼20.5 mg/dL (∼1.14 mmol/L) mean difference that was reported recently (32). The SD of venous to CGM responses were not different to duplicate meal SDs for Abbott and were higher for Dexcom, suggesting that the variance between CGM and venous measures was at least as high as the variance of duplicate meals within CGM. However, there were fewer pairs to calculate SD with the venous to CGM comparisons versus the within CGM comparison of duplicate meals and perhaps more repeated measures are required to reliably compare the variance between CGM and venous measures within each participant.

Our participants were representative of a generally healthy population across a wide range of body mass indexes, but without diabetes or other metabolic disease. Recent cross-sectional evidence using Medtronic devices suggests day-to-day reproducibility of CGM readings is lower in younger individuals (< 60 years) without prediabetes or type 2 diabetes (33). Intriguingly, we found a low mean bias of within-subject iAUCs in response to multiple pairs of duplicate meals suggesting that it may be possible to reliably estimate within-subject postprandial responses to the same meals provided that enough repeated measurements are made. Identifying the number of repeated postprandial CGM measurements, in response to the same meals within-participants, that is required to provide reliable personalized estimates, is a critical question for future research. Our results suggest that two measurements are too few even under highly standardized metabolic ward conditions.

In conclusion, our data suggest that personalized diet advice is unlikely to be reliable if it is based primarily on postprandial CGM measurements obtained using very few repeated measurements in adults without diabetes. Instead, precision nutrition requires more reliable methods involving aggregated repeated measurements.

## Data Availability

Deidentified individual data from consenting subjects will be made freely available upon final publication.

## Abbreviations

CGM: continuous glucose monitor
ICC: intra-class correlation coefficient

## Acknowledgements

This work was supported by the Intramural Research Program of the NIH, National Institute of Diabetes & Digestive & Kidney Diseases under award number 1ZIADK013037. We thank the nursing and nutrition staff at the NIH Metabolic Clinical Research Unit for their invaluable assistance with this study. We thank the study participants for their invaluable contribution.

## Author contributions

JG and KDH conceptualized and designed the research, AH, JAO, KM, JG, and KDH analyzed and interpreted the data, and critically reviewed, drafted, and approved the final manuscript.

## Online Supplemental File

### Supplemental Figures

**Supplemental Figure 1.**
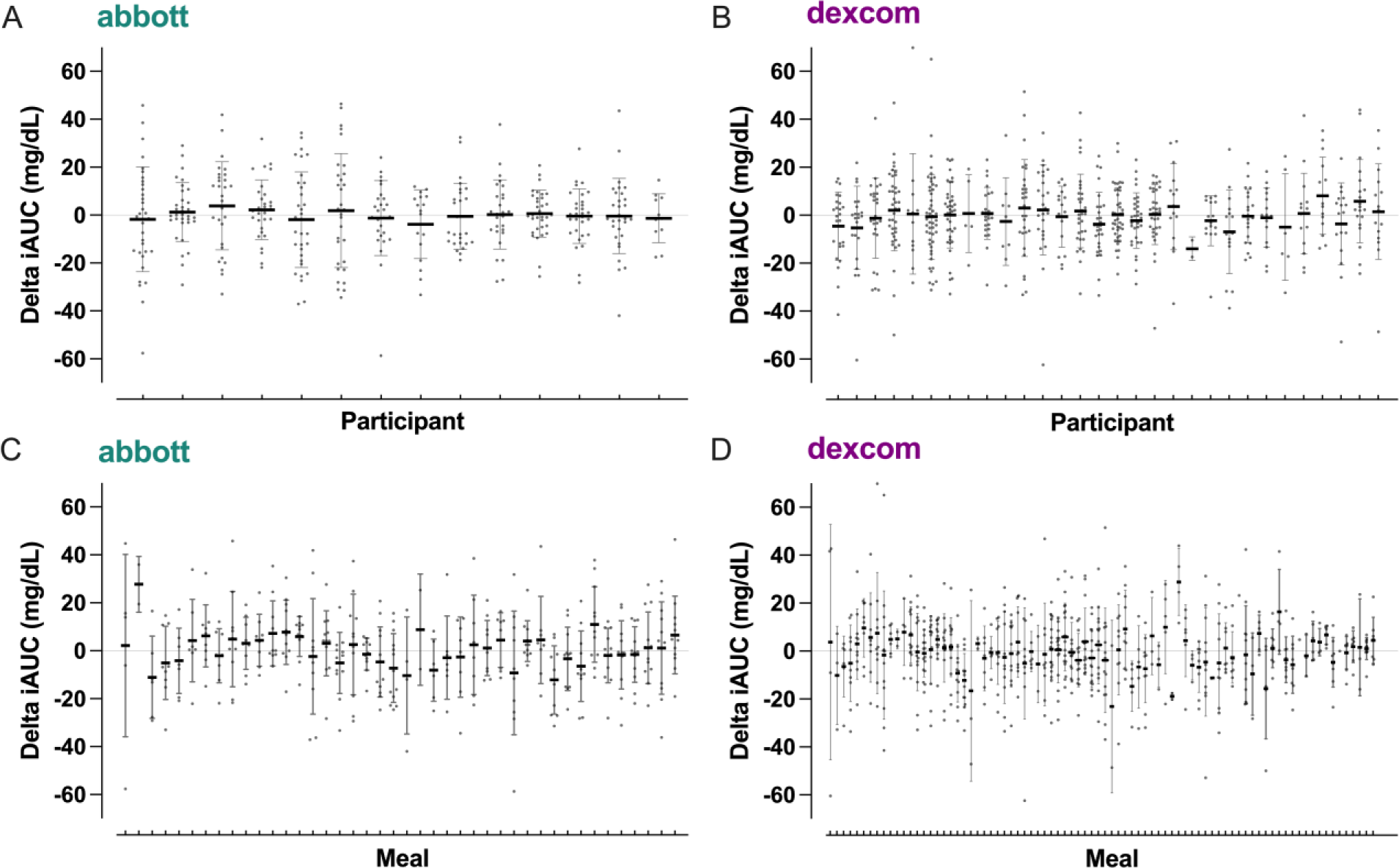
Mean ±SD difference and individual comparisons of duplicate meals organized by participant using A) Abbott and B) Dexcom devices. Each data point is the within-participant iAUC difference between duplicate meals. Mean ±SD and individual comparisons of duplicate meals ordered by meal pairing (across all participants) using C) Abbott and D) Dexcom CGMs. Each data point is a duplicate meal eaten in week 2 minus the same meal eaten in week 1 with data from all participants who consumed that meal (abbott has 42 total meals for comparison across the 14 days of rotating menu, 14 days x 3 meals; dexcom has 63 total meals for comparison across 21 days of rotating menu, 21 days x 3 meals).

**Supplemental Figure 2.**
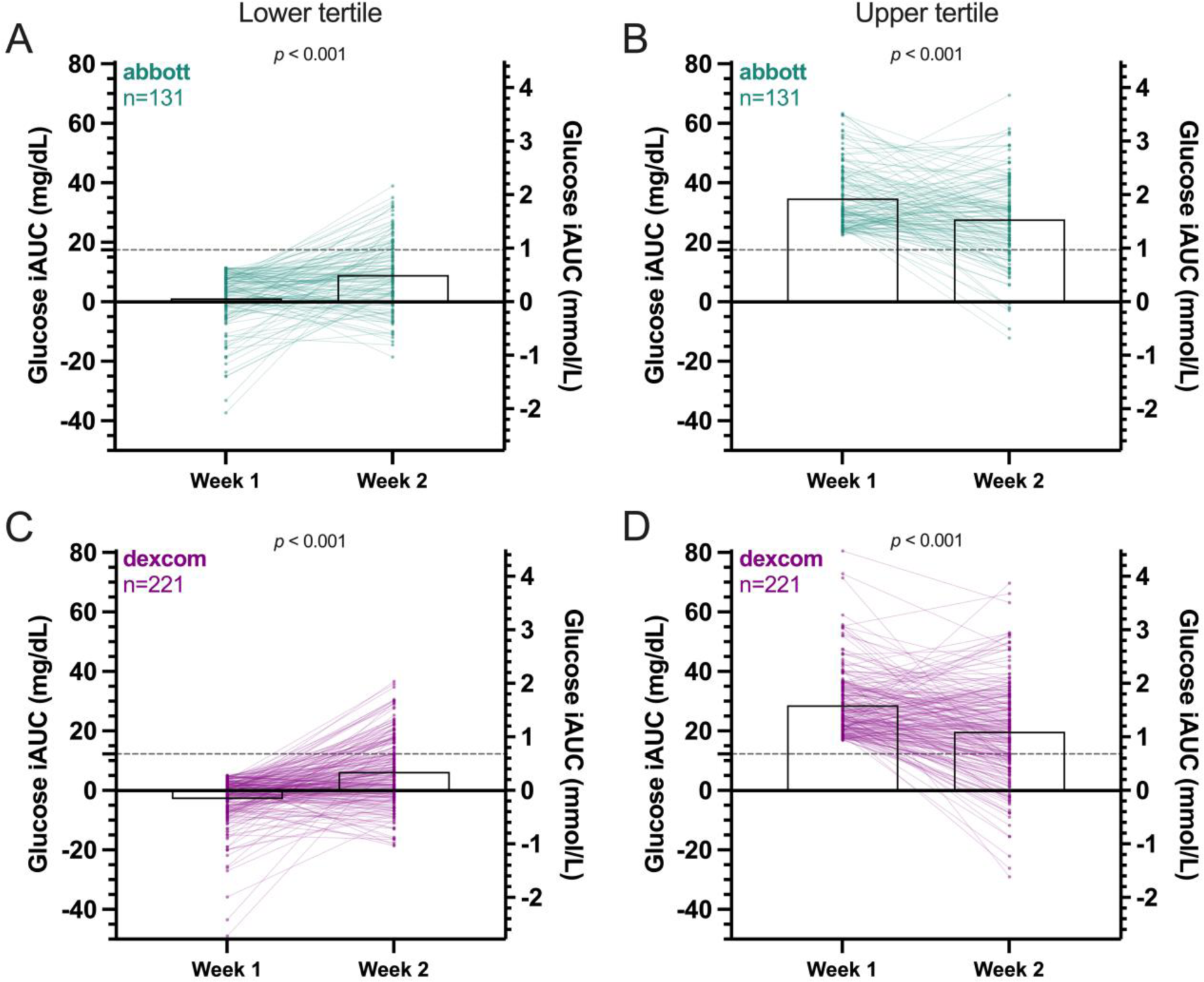
Mean and individual meal responses from lower tertile and upper tertile meals during week 1 and the corresponding comparisons in week 2. Lower tertile meals were significantly higher in week 2 for Abbott (**A**) and Dexcom (**C**) and upper tertile meals were significantly lower in week 2 for Abbott (**B**) and Dexcom (**D**). Dashed lines are mean of all presented meal responses across 2 weeks. iAUC = incremental area under the curve.

## References

1. Berry SE, Valdes AM, Drew DA, Asnicar F, Mazidi M, Wolf J, et al. Human postprandial responses to food and potential for precision nutrition. Nat Med. 2020;26(6):964–73. doi: 10.1038/s41591-020-0934-0.

2. Zeevi D, Korem T, Zmora N, Israeli D, Rothschild D, Weinberger A, et al. Personalized Nutrition by Prediction of Glycemic Responses. Cell. 2015;163(5):1079–94. doi: 10.1016/j.cell.2015.11.001.

3. Mendes-Soares H, Raveh-Sadka T, Azulay S, Edens K, Ben-Shlomo Y, Cohen Y, et al. Assessment of a Personalized Approach to Predicting Postprandial Glycemic Responses to Food Among Individuals Without Diabetes. JAMA Netw Open. 2019;2(2):e188102. doi: 10.1001/jamanetworkopen.2018.8102.

4. Hall KD, Ayuketah A, Brychta R, Cai HY, Cassimatis T, Chen KY, et al. Ultra-Processed Diets Cause Excess Calorie Intake and Weight Gain: An Inpatient Randomized Controlled Trial of Ad Libitum Food Intake (vol 30, pg 67, 2019). Cell Metabolism. 2019;30(1):226-. doi: 10.1016/j.cmet.2019.05.020.

5. Hall KD, Guo J, Courville AB, Boring J, Brychta R, Chen KY, et al. Effect of a plant-based, low-fat diet versus an animal-based, ketogenic diet on ad libitum energy intake. Nat Med. 2021;27(2):344–53. doi: 10.1038/s41591-020-01209-1.

6. Wolever TM. Effect of blood sampling schedule and method of calculating the area under the curve on validity and precision of glycaemic index values. The British journal of nutrition. 2004;91(2):295–301. doi: 10.1079/bjn20031054.

7. Bates D, Machler M, Bolker BM, Walker SC. Fitting Linear Mixed-Effects Models Using lme4. J Stat Softw. 2015;67(1):1–48. doi: DOI 10.18637/jss.v067.i01.

8. Koo TK, Li MY. A Guideline of Selecting and Reporting Intraclass Correlation Coefficients for Reliability Research. J Chiropr Med. 2016;15(2):155–63. doi: 10.1016/j.jcm.2016.02.012.

9. Bland JM, Altman DG. Agreement between methods of measurement with multiple observations per individual. J Biopharm Stat. 2007;17(4):571–82. doi: 10.1080/10543400701329422.

10. Jaklevic MC. Start-ups Tout Continuous Glucose Monitoring for People Without Diabetes. JAMA. 2021;325(21):2140–2. doi: 10.1001/jama.2021.3789.

11. Betts JA, Gonzalez JT. Personalised nutrition: What makes you so special? Nutr Bull. 2016;41(4):353–9. doi: 10.1111/nbu.12238.

12. Ben-Yacov O, Godneva A, Rein M, Shilo S, Kolobkov D, Koren N, et al. Personalized Postprandial Glucose Response-Targeting Diet Versus Mediterranean Diet for Glycemic Control in Prediabetes. Diabetes care. 2021;44(9):1980–91. doi: 10.2337/dc21-0162.

13. Kharmats AY, Popp C, Hu L, Berube L, Curran M, Wang C, et al. A randomized clinical trial comparing low-fat with precision nutrition-based diets for weight loss: impact on glycemic variability and HbA1c. Am J Clin Nutr. 2023. doi: 10.1016/j.ajcnut.2023.05.026.

14. Howard R, Guo J, Hall KD. Imprecision nutrition? Different simultaneous continuous glucose monitors provide discordant meal rankings for incremental postprandial glucose in subjects without diabetes. Am J Clin Nutr. 2020;112(4):1114–9. doi: 10.1093/ajcn/nqaa198.

15. Merino J, Linenberg I, Bermingham KM, Ganesh S, Bakker E, Delahanty LM, et al. Validity of continuous glucose monitoring for categorizing glycemic responses to diet: implications for use in personalized nutrition. Am J Clin Nutr. 2022;115(6):1569–76. doi: 10.1093/ajcn/nqac026.

16. Hall KD. Challenges of human nutrition research. Science. 2020;367(6484):1298-300. doi: 10.1126/science.aba3807.

17. Hall KD, Heymsfield SB, Kemnitz JW, Klein S, Schoeller DA, Speakman JR. Energy balance and its components: implications for body weight regulation. Am J Clin Nutr. 2012;95(4):989–94. doi: 10.3945/ajcn.112.036350.

18. Solomon TPJ, Tarry E, Hudson CO, Fitt AI, Laye MJ. Immediate post-breakfast physical activity improves interstitial postprandial glycemia: a comparison of different activity-meal timings. Pflugers Arch. 2020;472(2):271–80. doi: 10.1007/s00424-019-02300-4.

19. Coates AM, Cohen JN, Burr JF. Investigating sensor location on the effectiveness of continuous glucose monitoring during exercise in a non-diabetic population. Eur J Sport Sci. 2023;23(10):2109–17. doi: 10.1080/17461391.2023.2174452.

20. Arner P, Kriegholm E, Engfeldt P, Bolinder J. Adrenergic regulation of lipolysis in situ at rest and during exercise. The Journal of clinical investigation. 1990;85(3):893–8. doi: 10.1172/JCI114516.

21. Shankar SS, Vella A, Raymond RH, Staten MA, Calle RA, Bergman RN, et al. Standardized Mixed-Meal Tolerance and Arginine Stimulation Tests Provide Reproducible and Complementary Measures of beta-Cell Function: Results From the Foundation for the National Institutes of Health Biomarkers Consortium Investigative Series. Diabetes care. 2016;39(9):1602–13. doi: 10.2337/dc15-0931.

22. Shukla AP, Andono J, Touhamy SH, Casper A, Iliescu RG, Mauer E, Shan Zhu Y, Ludwig DS, Aronne LJ. Carbohydrate-last meal pattern lowers postprandial glucose and insulin excursions in type 2 diabetes. BMJ Open Diabetes Res Care. 2017;5(1):e000440. doi: 10.1136/bmjdrc-2017-000440.

23. Shukla AP, Dickison M, Coughlin N, Karan A, Mauer E, Truong W, et al. The impact of food order on postprandial glycaemic excursions in prediabetes. Diabetes Obes Metab. 2019;21(2):377–81. doi: 10.1111/dom.13503.

24. Shukla AP, Iliescu RG, Thomas CE, Aronne LJ. Food Order Has a Significant Impact on Postprandial Glucose and Insulin Levels. Diabetes care. 2015;38(7):e98–e9. doi: 10.2337/dc15-0429.

25. Trico D, Filice E, Trifiro S, Natali A. Manipulating the sequence of food ingestion improves glycemic control in type 2 diabetic patients under free-living conditions. Nutr Diabetes. 2016;6(8):e226. doi: 10.1038/nutd.2016.33.

26. Chen YC, Betts JA, Walhin JP, Thompson D. Adipose Tissue Responses to Breaking Sitting in Men and Women with Central Adiposity. Medicine and science in sports and exercise. 2018;50(10):2049–57. doi: 10.1249/MSS.0000000000001654.

27. Dunstan DW, Kingwell BA, Larsen R, Healy GN, Cerin E, Hamilton MT, et al. Breaking up prolonged sitting reduces postprandial glucose and insulin responses. Diabetes care. 2012;35(5):976–83. doi: 10.2337/dc11-1931.

28. Peddie MC, Bone JL, Rehrer NJ, Skeaff CM, Gray AR, Perry TL. Breaking prolonged sitting reduces postprandial glycemia in healthy, normal-weight adults: a randomized crossover trial. Am J Clin Nutr. 2013;98(2):358–66. doi: 10.3945/ajcn.112.051763.

29. Pettit-Mee RJ, Ready ST, Padilla J, Kanaley JA. Leg Fidgeting During Prolonged Sitting Improves Postprandial Glycemic Control in People with Obesity. Obesity. 2021;29(7):1146–54. doi: 10.1002/oby.23173.

30. Tsereteli N, Vallat R, Fernandez-Tajes J, Delahanty LM, Ordovas JM, Drew DA, et al. Impact of insufficient sleep on dysregulated blood glucose control under standardised meal conditions. Diabetologia. 2022;65(2):356–65. doi: 10.1007/s00125-021-05608-y.

31. Basu A, Dube S, Slama M, Errazuriz I, Amezcua JC, Kudva YC, Peyser T, Carter RE, Cobelli C, Basu R. Time lag of glucose from intravascular to interstitial compartment in humans. Diabetes. 2013;62(12):4083–7. doi: 10.2337/db13-1132.

32. Jin Z, Thackray AE, King JA, Deighton K, Davies MJ, Stensel DJ. Analytical Performance of the Factory-Calibrated Flash Glucose Monitoring System FreeStyle Libre2(TM) in Healthy Women. Sensors (Basel). 2023;23(17). doi: 10.3390/s23177417.

33. Matabuena M, Pazos-Couselo M, Alonso-Sampedro M, Fernandez-Merino C, Gonzalez-Quintela A, Gude F. Reproducibility of continuous glucose monitoring results under real-life conditions in an adult population: a functional data analysis. Sci Rep. 2023;13(1):13987. doi: 10.1038/s41598-023-40949-1.

